# Antenatal Care Dropout and Associated Factors in Ethiopia: A Systematic Review and Meta-Analysis

**DOI:** 10.1101/2023.01.19.23284767

**Authors:** Gizaw Sisay, Tsion Mulat

## Abstract

**Background:** Antenatal care during pregnancy is one of the most important strategies for improving maternal and newborn health and preventing maternal and newborn mortality and morbidity. The prevalence and predictors of antenatal care dropout in Ethiopia were studied, and the results were inconsistent and showed considerable variation. Hence, this meta-analysis aimed at estimating the overall prevalence of antenatal care dropout and its associated factors in Ethiopia.

**Methods:** A comprehensive search of studies published before December 30, 2022, was explored by using distinct international databases such as (PubMed, DOJA, Embase, Cochrane library, African journals online, Google scholar, web of science and the institutional repository of Ethiopian universities were used to search relevant studies. Data were extracted using Microsoft Excel, and analysis was performed using STATA version 16. A random-effects model were used to estimate the overall prevalence of antenatal care drop-out and odd ratio for determinant factors. *I*^2^ Test-statistics for to assessing heterogeneity and Egger’s test for assessing publication bias were used.

**Results:** A total of seven studies were included for this systematic review and meta-analysis with of 11839 study participants. The overall pooled prevalence of antenatal dropout in Ethiopia was found to be 41.37% with 95% CI: (35.04, 47.70). Distance from the health care facility (AOR = 2.55, 95% CI = 1.79, 3.31), pregnancy complication signs (AOR = 2.88, 95% CI= 2.41, 3.66), place of residence (AOR= 1.59, 95% CI = 1.31, 1.87), educational level (AOR=1.79, 95%CI = 1.37, 2.21), age group(30-49) (AOR=(AOR = 0.57, 95% CI = 0.26, 0.88) were significantly associated with antenatal care dropout.

**Conclusion:** Based on this systematic review and meta-analysis, 41% of Ethiopian women dropped out of antenatal care visits before the minimum recommended visit (four times). Hence, to reduce the number of ANC dropouts it is important to counsel and educate women at their first prenatal care. Issues of urban-rural disparity and locations identified as hotspots for incomplete ANC visits require that further attention.

## Introduction

Antenatal care plays a great role for the pregnant women to improve of health of the women in getting ready for labor and in recognizing the warning signs of pregnancy and childbirth. Dropping out of antenatal care is defined as ceasing to receive the service before at least four antenatal care visits [1]. Antenatal care (ANC) is one of the approaches to reduce newborn mortality and maintain maternal health by guiding women to access more maternal health services and prepare for their newborn [2].

In 2017, around 810 women died every day from preventable causes related to pregnancy and childbirth. 94% of all maternal deaths occur in low- and lower-middle-income countries. Despite an apparent global improvement, between 2000 and 2017 maternal mortality dropped by about 38% [3].

In 2016, at the beginning of the Sustainable Development Goals (SDGs) era, pregnancy-related morbidity and mortality rates are still too high. While substantial progress has been made, countries need to consolidate and increase these advances, and to expand their agendas to go beyond survival, with a view to maximizing the health and potential of their populations. The sustainable development Goals target a global maternal mortality ratio not greater than 70 maternal deaths per 100 000 live births by 2030 [4].

Women in low-resource setting countries including Ethiopia were prone to high rates of maternity care dropout as well as other maternal continuum of care issues [5]. In Ethiopia, only 32% of women had four or more ANC visits and 64% of women had at least one ANC visit, while about 32% of women did not receive ANC at all throughout the duration of their pregnancy [6].

Antenatal care visit dropout was high in low-income countries. The sub-Saharan region was one of the African regions with a high dropout rate from antenatal care. In the region, about 30% of pregnant women among all births in the region did not use the services [7].

Although maternal health care utilization, including ANC has improved in Ethiopia, the majority of women do not attend the minimum recommended number of ANC visits required [8]. Low health care utilization is caused by a variety of factors, but it is clear that poor quality of services and poor treatment by providers are among the reasons why many women do not access newborn and maternal health care services [9].

In Ethiopia different studies conducted on the prevalence of ANC dropout and lies between 33.39% [10] to 56.49% [11]. Different factors affect the dropout of ANC services utilization were Being far distance, Not developing a pregnancy danger sign, Not satisfied in ANC service, Having no formal education, employment status, marital status, Didn’t informed about institutional delivery, place of residence, age group, current pregnancy planned/supported, presence of professional advice during ANC visit [5, 6, 10-15].

As Health Sector Transformation Plan 2 (HSTP 2) come to the end, which aimed to promote maternity care, with an emphasis on ANC to improve maternal and newborn health outcomes [16]. Therefore, it is important to the assesse the overall magnitude of antenatal care dropout and its determinant factors to produce evidences for decision making for planners and program evaluators. Even though there have been numerous studies conducted on the prevalence and determinant factors of ANC dropout in Ethiopia, the results have been inconsistent and varied.

## Methods

This systematic review and meta-analysis was conducted according to the guidelines of the Preferred Reporting Items for Systematic Reviews and Meta-Analyses (PRISMA) 2020 [17]. The PEO (Population, Exposure of Interest, Outcome) technique from the 2017 JBI Guideline was used to define the inclusion and exclusion criteria [18].

### Search strategies

A comprehensive search of studies were conducted using distinct databases such as (PubMed, DOJA, Embase, science direct, Cochrane library, African journals online, Google scholar, and web of science). At the beginning, studies were comprehensively searched by using the full title (“The prevalence and associated factors of antenatal care service dropout in Ethiopia”) and keywords (“prevalence,” “magnitude,” “incomplete,” “antenatal care dropout,” “determinant factors,” “associated factors,” “predictors,” “in Ethiopia”). Boolean operators “OR” or “AND” were used in combination or independently to connect these keywords and to establish the search terms. Besides, reference lists of all included studies were considered to find other missed studies.

Furthermore, institutional repository from Mekelle, Jima, Addis Abeba, and Harameya University was used to search important literature, specifically unpublished thesis reports, doctoral theses. Then Endnote v-X7.2 reference management software was used to combine search results from databases and remove duplicate articles initially.

#### Criteria’s of Eligibility

To establish the inclusion and exclusion criteria for this systematic review and meta-analysis, the authors used the PICO technique, which mainly involves CoCoPop (Condition, Context, and Population) questions for the prevalence studies.

### Inclusion Criteria

▪ **Study area:** Studies conducted in Ethiopia.
▪ **Study setting:** studies conducted in rural or urban settings and at health institutions or community-based settings.
▪ **Study design:** Cross-sectional, cohort, and case-control studies which reports either the proportion or prevalence or magnitude and associated or determinants factors or predictors of antenatal care dropout.
▪ **Time:** No limitation was made for the time of publication and all studies conducted in Ethiopia were included.

### Exclusion criteria

Qualitative studies and studies that have a different outcome of interest were excluded. Besides this, studies with different operational definitions and measurements of antenatal care dropout were excluded from this systematic review and meta-analysis.

### Data extraction

Two authors (G.S and T.M) separately extracted the data from the included studies using a standardized data extraction Microsoft Excel spreadsheet. The spreadsheet includes the first author’s name, publication year, study year, study design, study area, study setup, sample size, response rate, and the proportion of antenatal care dropout. When there was any discrepancy between the extractors, the differences were resolved through discussion and then re-evaluation of the studies were conducted.

### Quality assessment of the studies

Joanna Briggs Institute (JBI) Critical appraisal checklist was used to assess the methodological quality of the included studies [19]. The two authors individually ranked the quality of the included studies. There are nine parameters in the tool as follows:

1. Was the sampling frame appropriate to address the target population?
2. Were study participants sampled appropriately?
3. Was the sample size adequate?
4. Were the study subjects and the setting described in detail?
5. Was the data analysis conducted with sufficient coverage of the identified sample?
6. Were valid methods used for the identification of the condition?
7. Was the condition measured in a standard, reliable way for all participants?
8. Was there appropriate statistical analysis?
9. Was the response rate adequate, and if not, was the low response rate managed appropriately?

Each item was evaluated as either low or high risk of bias. A composite quality index was grouped and the risk of bias was ranked as low (0–2), moderate (3 or 4), or high (>5). Articles with low and moderate risks of bias were considered for this systematic review and meta-analysis.

### Measurement of the outcome of interest

This systematic review and meta-analysis had two outcomes (antenatal care services dropout and its associated factor) in Ethiopia. Antenatal care services dropout was considered when the pregnant mothers did not complete full recommended visit during their pregnancy (a minimum of 4 visits for normal pregnancy) [10-13, 15, 20].

## Statistical methods and analysis

The extracted data from Microsoft Excel spreadsheet were imported to STATA™ 16 Statistical software for analysis. To check the presence of statistical heterogeneity among the included studies I-squared statistic (*I*^2^) statistic on forest plots were computed were used. As a result, significant heterogeneity [*I*^2^ = 97.6%, p-value 0.001] was found. Therefore, a random-effects model using the DerSimonian-Laird method was used to estimate the pooled prevalence of ANC dropout and the corresponding 95% CI presented on a forest plot. The AORs from the include studies were extracted, with their corresponding 95% CIs. A random fixed effects model was also used to compute the pooled ORs with 95% CIs, and displayed on a forest plots.

### Subgroup analyses and heterogeneity

To identify potential sources of heterogeneity between the included studies, subgroup analyses were conducted based on geographic regions and year the study conducted. Meta-regression was done to assess the presence of significant heterogeneity.

### Publication bias

The presence of publication bias was assessed by using Statistical methods such as Egger’s and Begg’s tests, and the results of egger’s regression test indicated that no publication bias between the included studies (p-value > 0.05). A p-value of 0.233 and 0.114 for Begg’s and Egger’s tests, respectively, implied that a small-study effect was less likely.

### Sensitivity analysis

To assess the effect of a single study on the overall prevalence of ANC dropout, sensitivity analysis was done using a random-effects model.

## Results

Search of studies from different international database and the institutional repository of Ethiopian universities were carried out. A total of 265 published and unpublished studies were identified. The Endnote reference manager software was used to screen studies that was obtained and the duplicate studies were removed. Then, following the removal of 178 research based on title and abstract screening, 19 complete papers were obtained. Finally, this study analysis contained seven studies that met the inclusion criteria (Fig 1).

**Fig 1.**
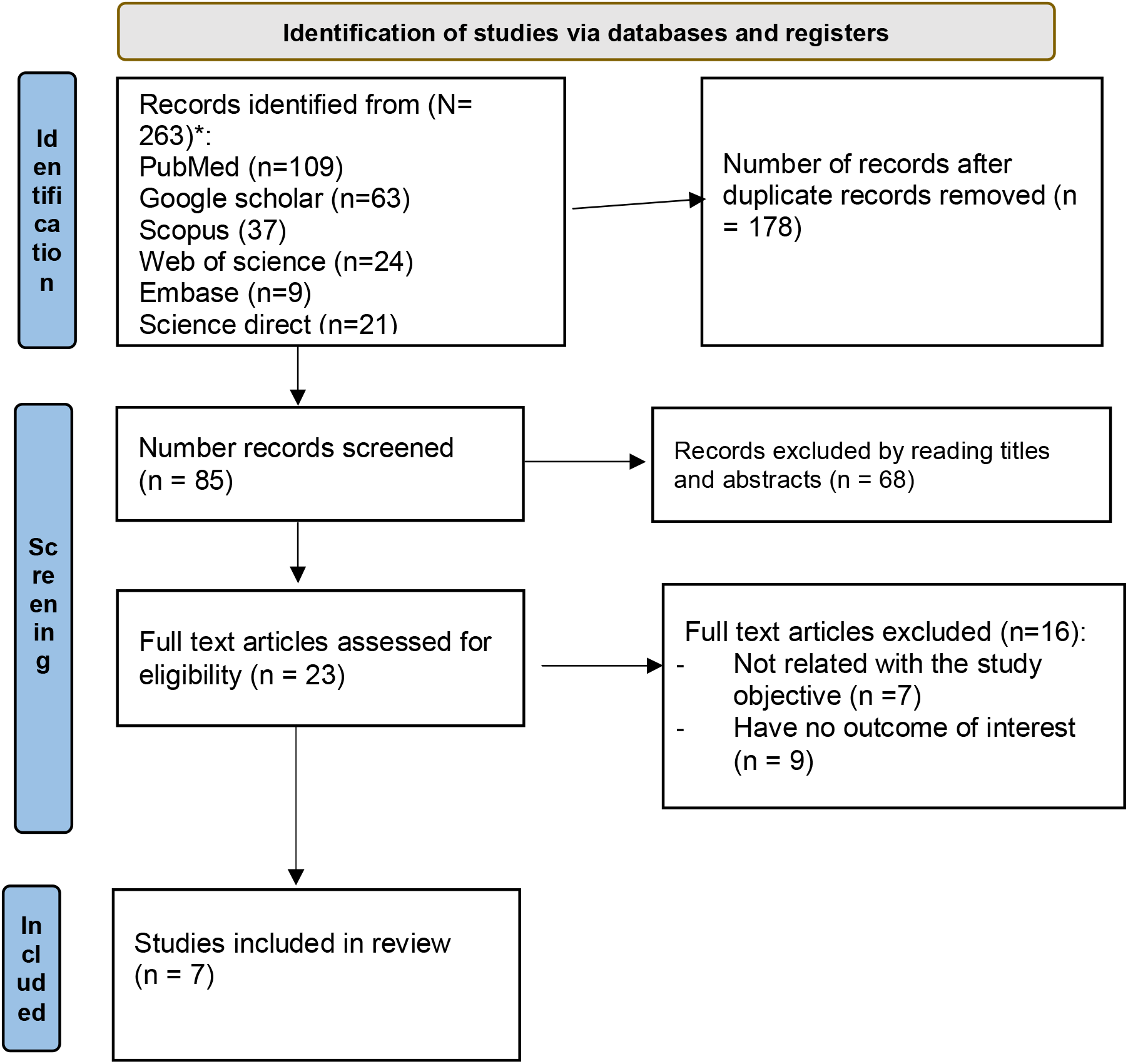
PRISMA 2020 flow diagram describing the selection of studies for systematic review and meta-analysis

### Characteristics of selected studies

A total of seven studies were included to assess the pooled prevalence of antenatal care service drop-out and its associated factors in Ethiopia [10-15]. A total of 11839 pregnant women were considered in this meta-analysis. In each studies, the methods of data collection were through face-to-face interview using a pre-tested, interviewer-administered questionnaire. The included studies publication year ranged from 2020 to 2022. The two from Amhara [12, 13], one from Tigray [10] and one from Oromia [15] regions and three of them were nationwide studies[5, 11, 14] based on Ethiopian demographic and health survey data. All studies were cross-sectional studies and have a low risk of bias (Table 1).

**Table 1.**
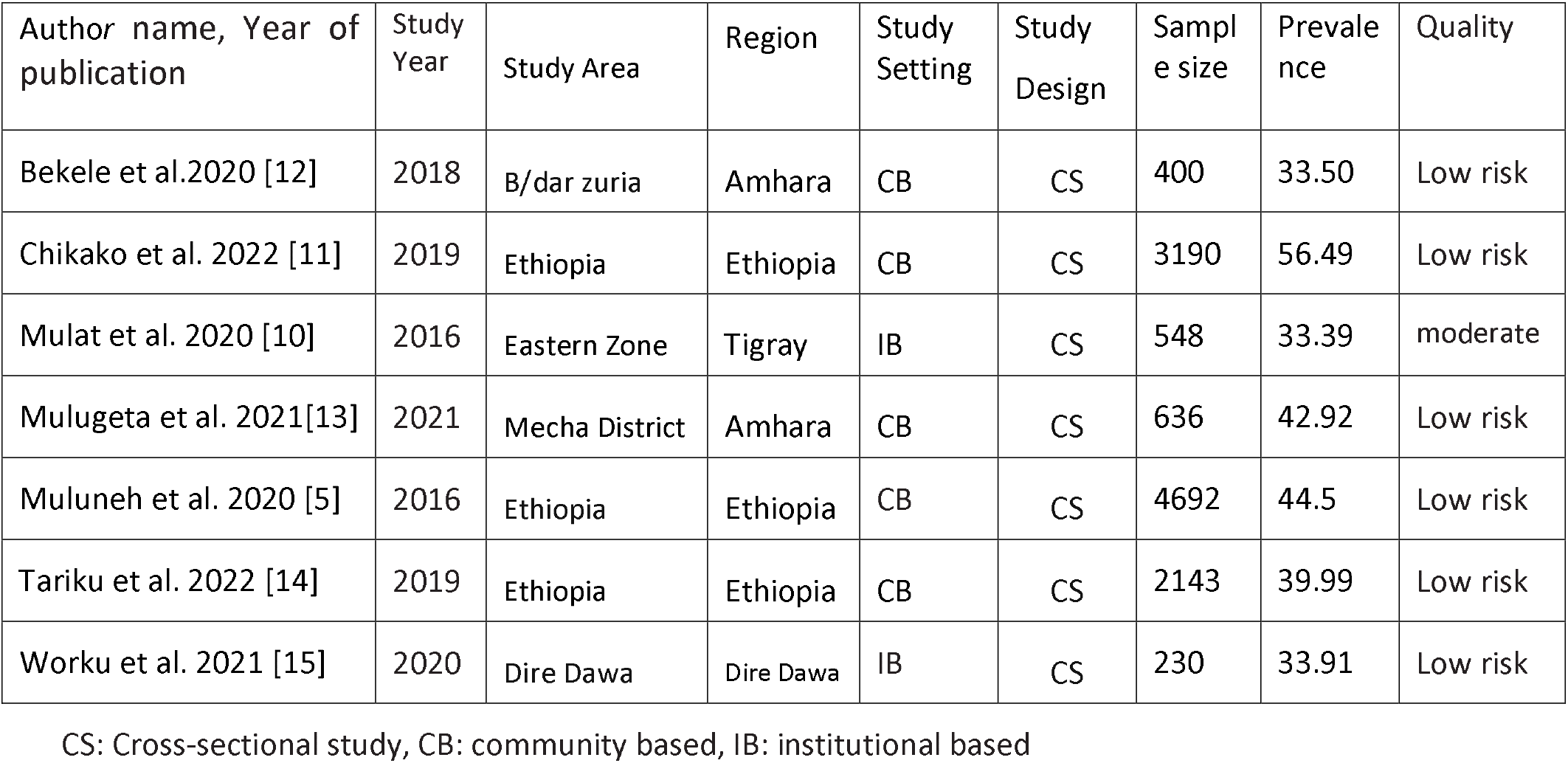
Shows a descriptive summary of seven studies included in this systematic review and meta-analysis.

| Author name, Year of publication | Study Year | Study Area | Region | Study Setting | Study Design | Sample size | Prevalence | Quality |
| --- | --- | --- | --- | --- | --- | --- | --- | --- |
| Bekele et al.2020 [12] | 2018 | B/dar zuria | Amhara | CB | CS | 400 | 33.50 | Low risk |
| Chikako et al. 2022 [11] | 2019 | Ethiopia | Ethiopia | CB | CS | 3190 | 56.49 | Low risk |
| Mulat et al. 2020 [10] | 2016 | Eastern Zone | Tigray | IB | CS | 548 | 33.39 | moderate |
| Mulugeta et al. 2021[13] | 2021 | Mecha District | Amhara | CB | CS | 636 | 42.92 | Low risk |
| Muluneh et al. 2020 [5] | 2016 | Ethiopia | Ethiopia | CB | CS | 4692 | 44.5 | Low risk |
| Tariku et al. 2022 [14] | 2019 | Ethiopia | Ethiopia | CB | CS | 2143 | 39.99 | Low risk |
| Worku et al. 2021 [15] | 2020 | Dire Dawa | Dire Dawa | IB | CS | 230 | 33.91 | Low risk |

CS: Cross-sectional study, CB: community based, IB: institutional based

### The pooled prevalence of antenatal care service dropout

The overall pooled prevalence of antenatal care drop-out in Ethiopia was found to be 41.37% (95% CI =35.04, 47.70) (Fig 2). The *I*^2^ test was computed to look at study heterogeneity, and the results showed that there was a substantial amount of variation between the included studies (*I*^2^=97.6%, P-value = 0.00).

**Fig 2.**
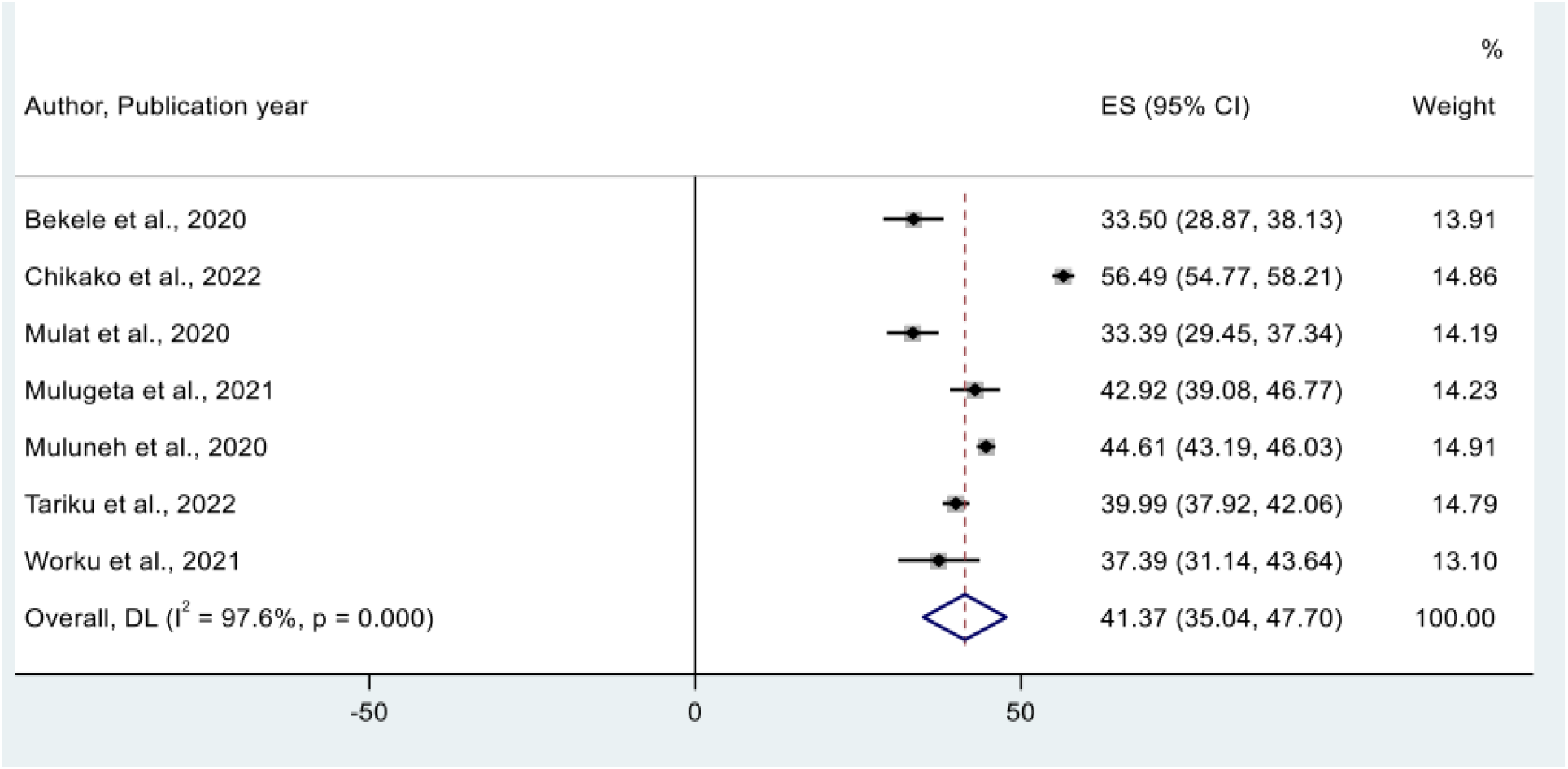
Forest plot showing the pooled estimate of antenatal care dropout in Ethiopia.

### Subgroup Analysis

To identify the source of heterogeneity for the pooled prevalence of ANC drop-out, subgroup analysis were done by place of region and year of the study conducted. The pooled prevalence of ANC dropout was significantly higher among studies conducted at national level (47.04%, 95% CI=37.86, 56.22). However, study conducted in Tigray region had the lowest prevalence of antenatal care service dropout 33.39% (95% CI: 29.45, 37.34) (Fig 3).

**Fig 3.**
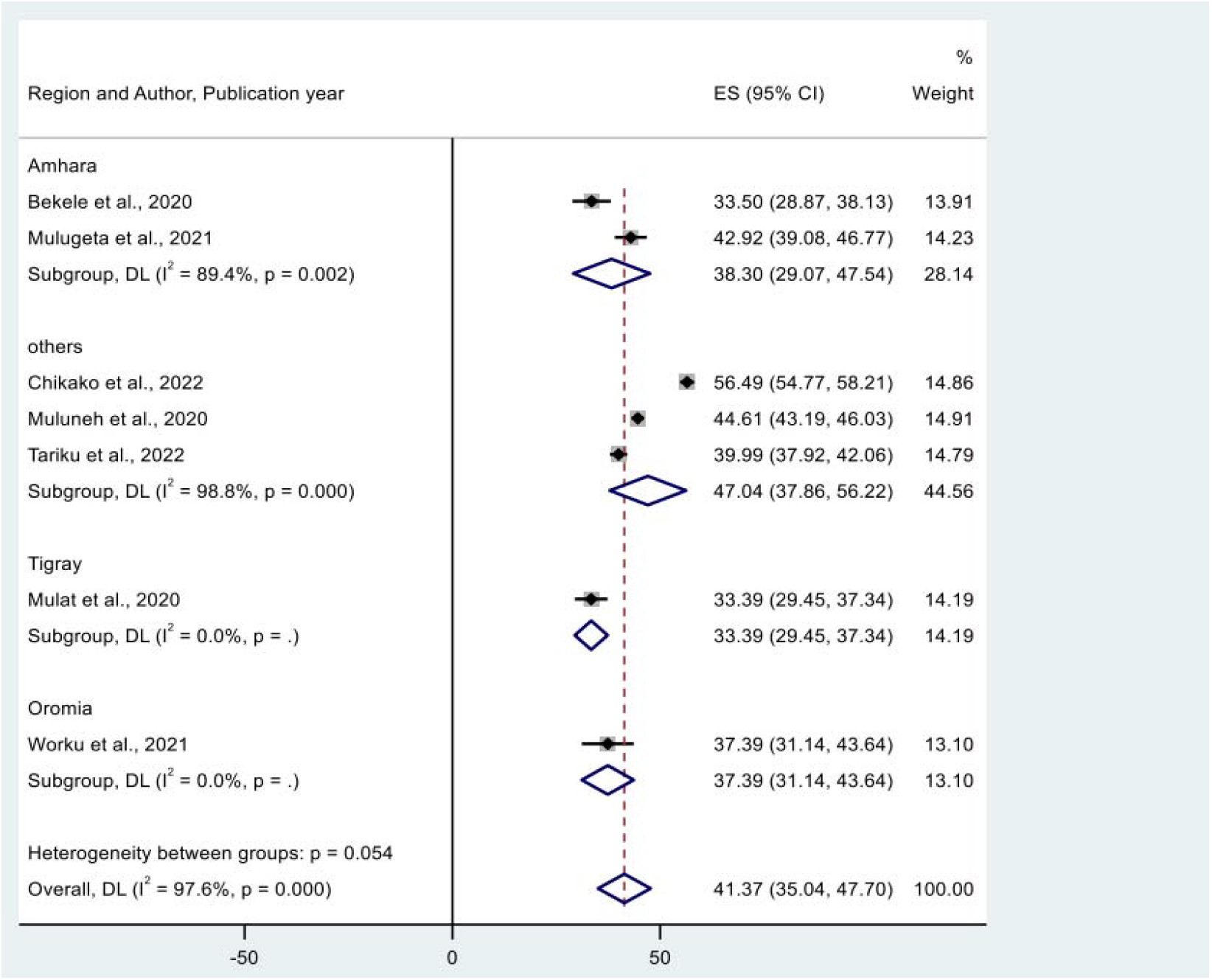
Sub-group analysis for the pooled prevalence of ANC dropout by geographical regions of Ethiopia.

Additionally, we carried out a subgroup analysis according to the year the studies were done. As a result, the pooled prevalence of antenatal care dropout was higher for those studies conducted in 2019 and after (44.34%, 95% CI= 34.05, 54.65 as compared to for those studies conducted before 2018 (37.35%, 95% CI=28.60, 46.10) (Fig 4).

**Fig 4.**
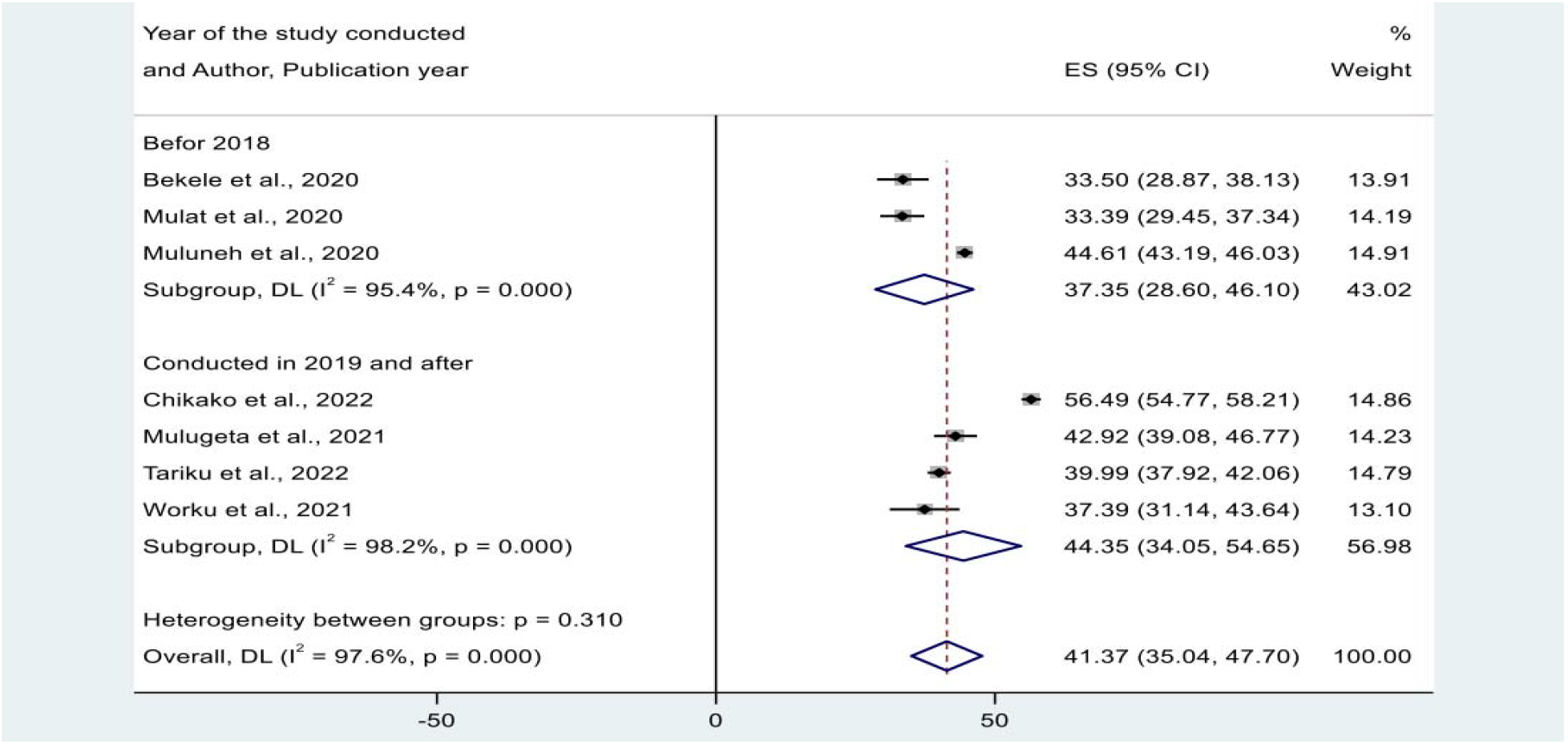
Sub-group analysis for the pooled prevalence of ANC dropout by year of the study conducted In Ethiopia.

To assess for underlying heterogeneity, meta-regression analysis were computed by using publication year and sample size, however there was statistically insignificant for underlying heterogeneity (p-value=0.161) and (p=0.111), respectively (Table 2)

**Table 2.** Meta-regression analysis based on sample size and year of publication.

| Possible source of heterogeneity | Coef. | Std. Err. | P-value |
| --- | --- | --- | --- |
| Publication year | 5.323 | 3.24 | 0.161 |
| Sample size | 0.0031 | 0.0016 | 0.111 |

#### Small study effect

By Using Egger’s regression test, publication biases among the included studies were evaluated, and the results showed that there was no small-study effects among the included studies as the non-statistical significant result (p-value = 0.114).

#### Sensitivity analysis

A sensitivity analysis with a random-effects model was done to examine any outliers on overall estimate of the pooled prevalence of antenatal care dropout and it showed that there was no single study influence (Fig 5).

**Fig 5.**
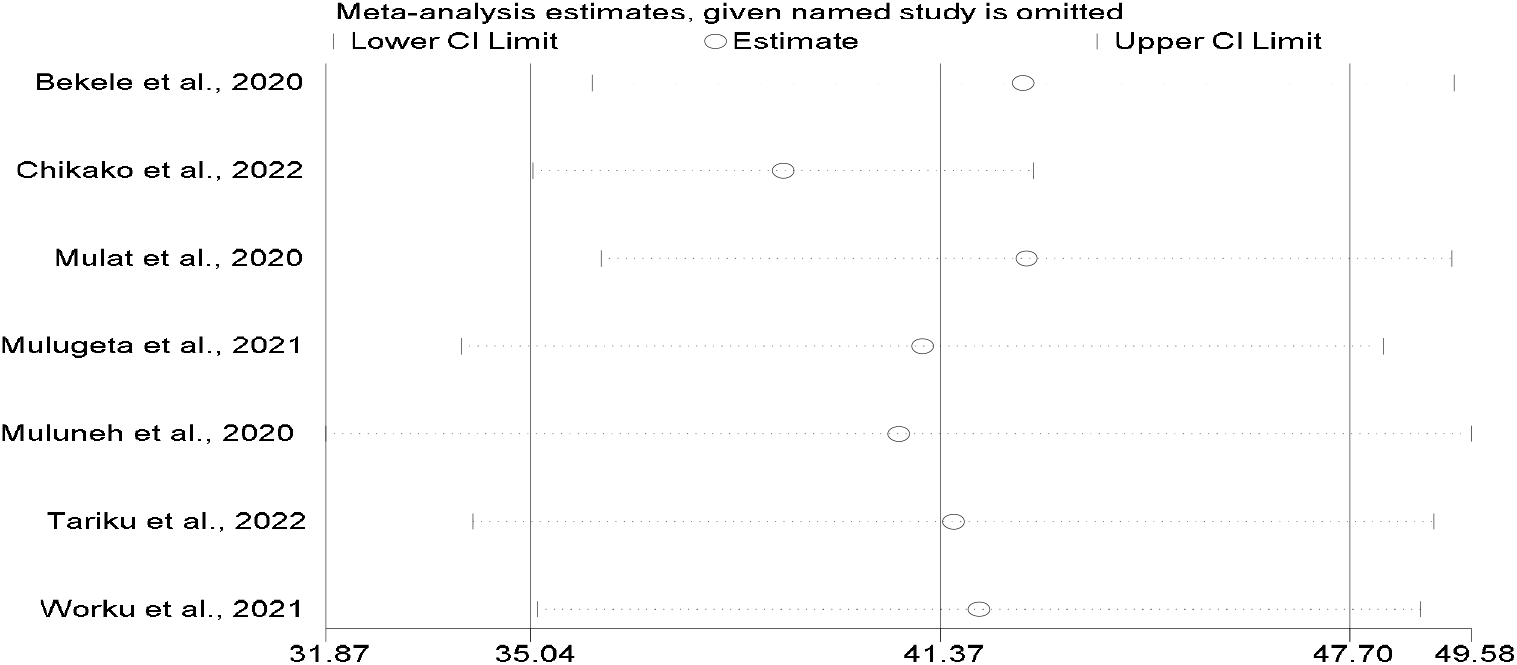
Result of sensitivity analysis of ANC dropout in Ethiopia.

### Predictors of Antenatal care Dropout in Ethiopia

To identify significant determinant factors of antenatal care dropout, twenty one variables were extracted from the included studies. Finally fife variables (distance from the health care facility, educational level, place residence, pregnancy complication signs and age group) were identified as significant determinants of ANC dropout.

The influence of distance from the health care facility on ANC dropout was assessed by using the findings of four studies [10-13]. In this meta-analysis, women who travel more than 1h to reach the health institution was 2.55 times more likely to have antenatal care dropout than their counterparts to [AOR = 2.93, 95% CI: 2.75, 3.11] (Fig 6).

**Fig 6.**
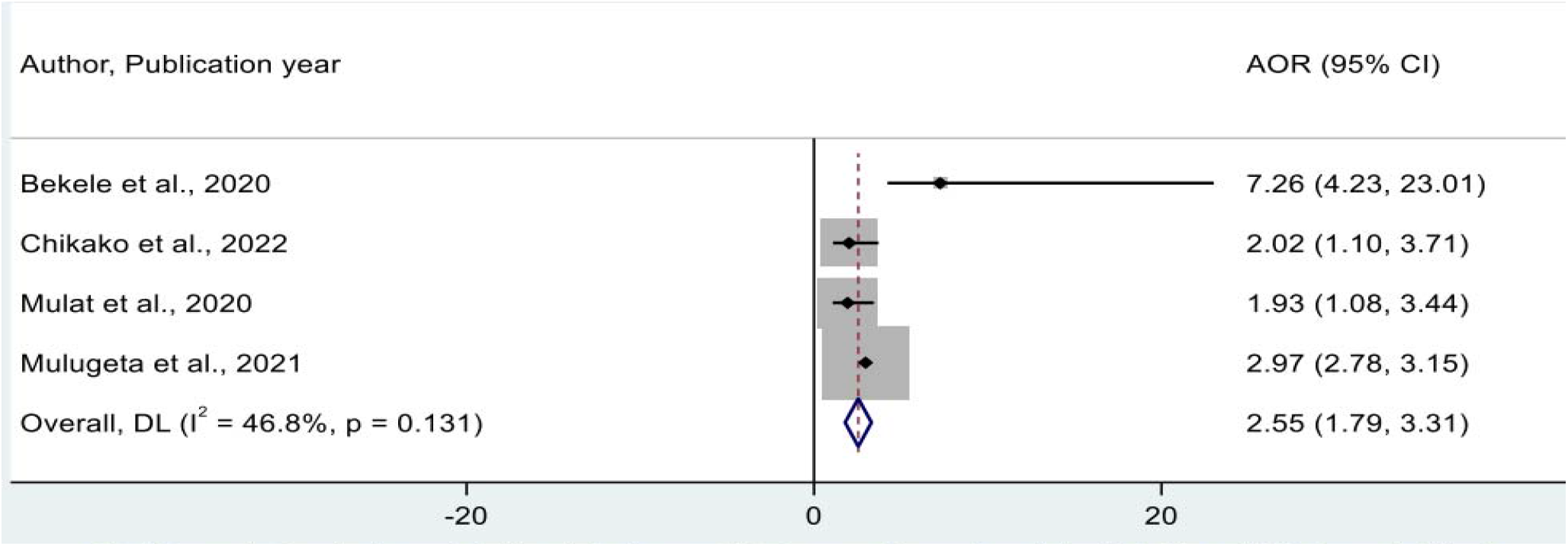
Forest plot showing the pooled odds ratio for the association between distance from the health facility and ANC dropout in Ethiopia.

To examine the association between place of residence and antenatal care dropout two studies were used [11, 14]. In this study pregnant women from rural areas were about 1.6 times more likely to have ANC dropout compared to women from urban areas counterparts (AOR= 1.59, 95% CI= 1.31, 1.87) (Fig. 7).

**Fig 7.**
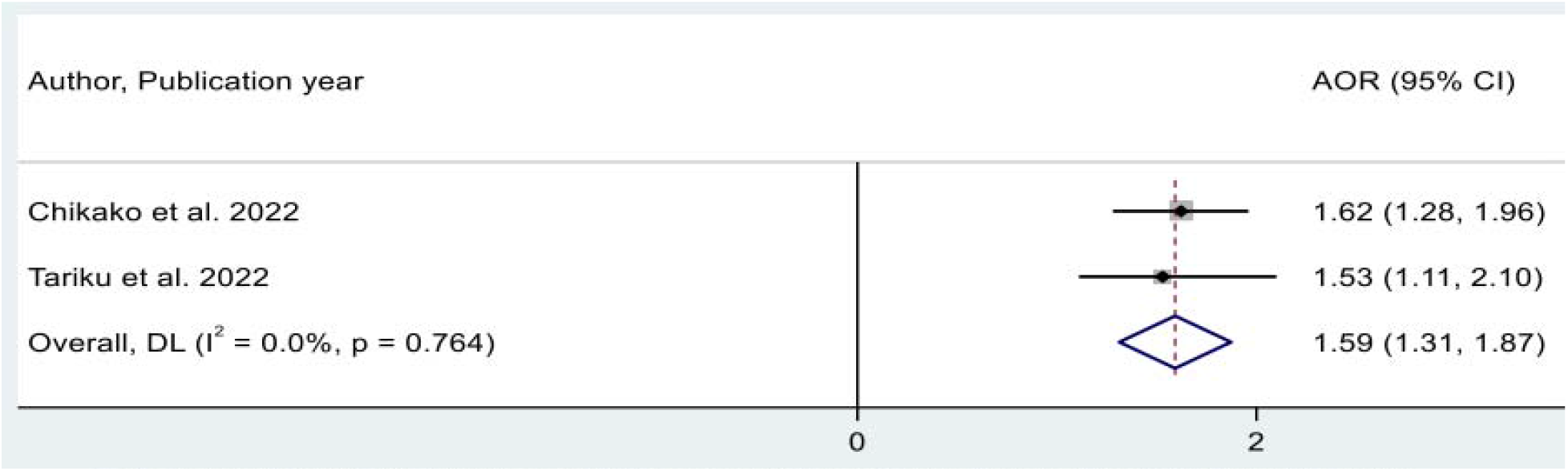
Forest plot showing the pooled estimate for the association between place of residence and ANC dropout in Ethiopia.

Additionally, two studies [11, 12] were used to assess the pooled odd ratio for the association between pregnancy complication signs and antenatal care dropout. Accordingly, pregnant women who had no pregnancy complication signs were 2.96 times more likely to dropout from ANC compared to the counterparts (AOR=2.96, 95% CI= 2.77, 3.16) (Fig 8).

**Fig 8.**
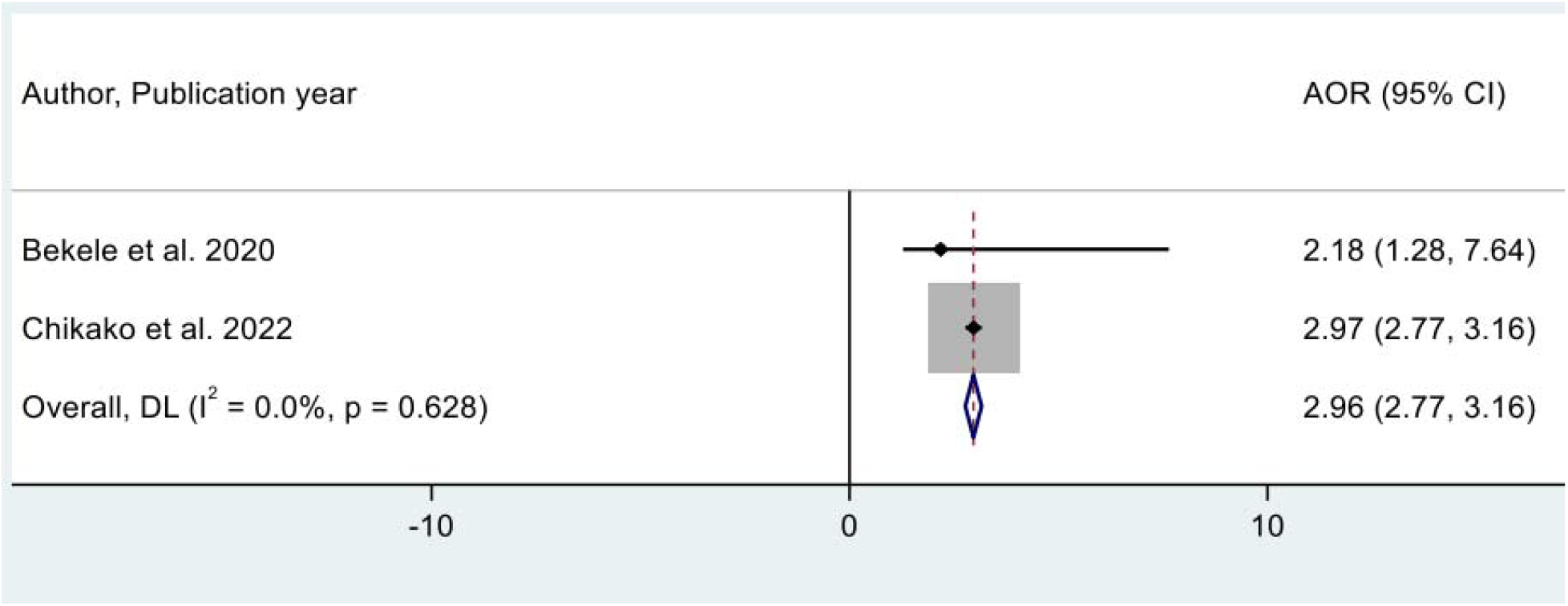
Forest plot showing the pooled odds ratio for the association between pregnancy complication signs and ANC dropout in Ethiopia.

Furthermore, in this meta-analysis the pooled effects of educational level were assessed by two studied [10, 14]. The results also indicate that women who had no educational level were about 1.59 more likely to dropout from ANC compared to those women who have secondary and above educational level (AOR=1.59, 95%CI= 1.31, 1.87) (Fig 9).

**Fig 9.**
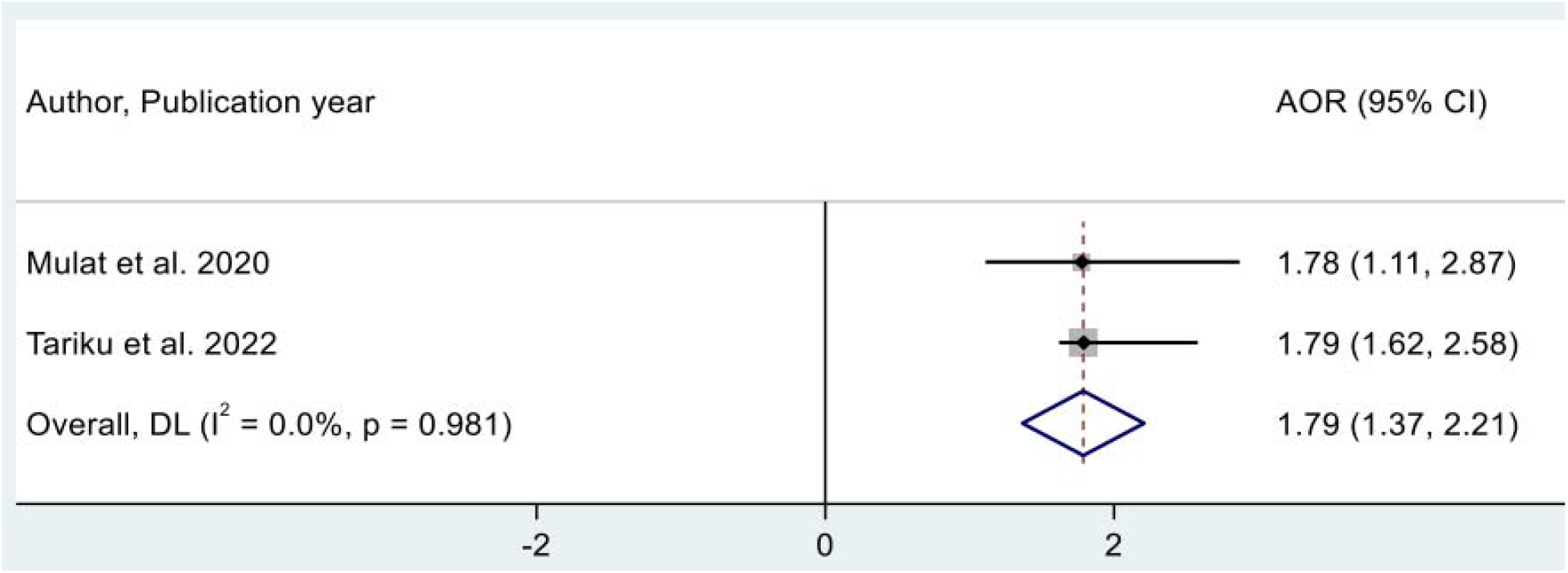
Forest plot showing the pooled odds ratio for the association between educational level and ANC dropout in Ethiopia.

Finally, the effect of age group on the antenatal care dropout was assessed by using two studies [14, 15], and the likelihoods of antenatal care services dropout were 43% (AOR = 0.57, 95% CI: 0.26, 0.88) lower among the pregnant women aged 30-49 years as compared with the pregnant women aged 15-29 years (Fig 10).

**Fig 10.**
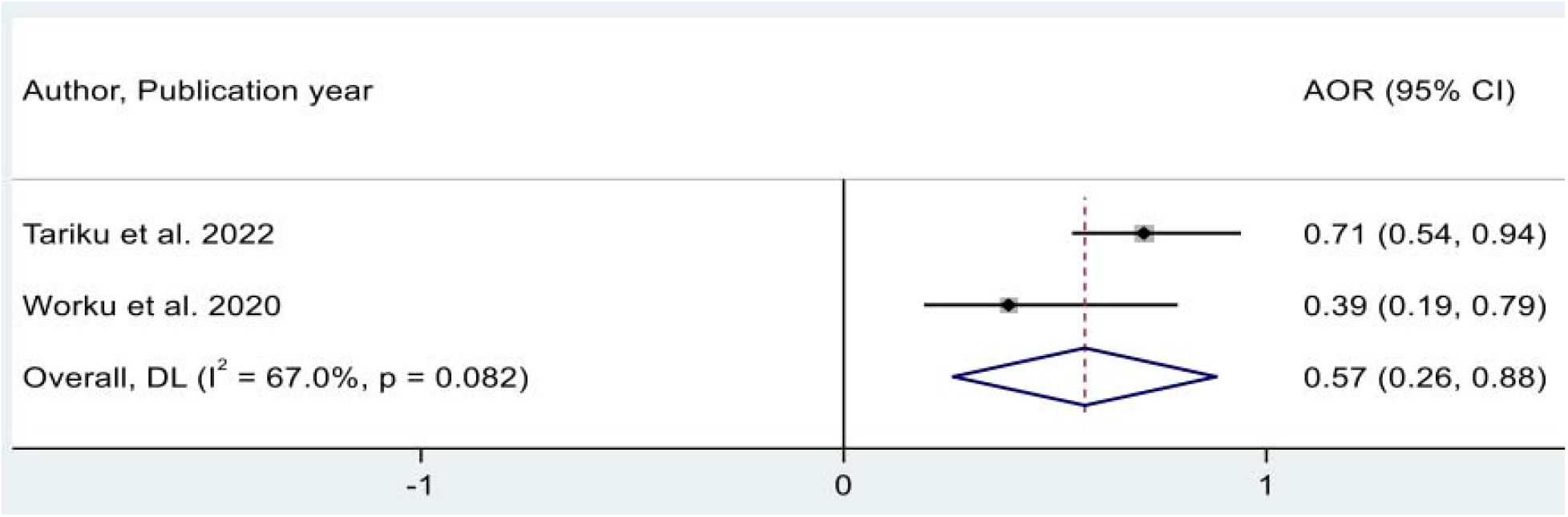
Forest plot showing the association between age group and ANC service dropout.

## Discussion

This meta-analysis aimed to assess the pooled prevalence and determinant factors of antenatal care service dropout in Ethiopia. Accordingly, the overall prevalence of the antenatal care dropout among women on ANC follow-up in Ethiopia was found to be 41.37% with 95% CI (35.04, 47.70). Despite the absence of systematic reviews and meta-analyses undertaken in Ethiopia or elsewhere, we contrasted the current pooled prevalence with a number of primary research conducted aboard. As a result, the prevalence of antenatal care dropout was higher as compared to the results of primary studies conducted in Nigeria 38.1% [21] and Kenya 29.2% [22]. This variation might be due to the fact that health care providers play a crucial role in Ethiopia by raising awareness of ANC through counseling and education. However, the result of this study was also lower than primary studies conducted in Uganda 55.8% [23] and Nepal 50.47% [24]. This variation could be due to differences in accessibility of health facilities, quality of maternal health services and lack of awareness about antenatal care service; as noted in most of the studies included in this systematic review and meta-analysis [5, 13, 14].

In this review from subgroup analysis was done by geographical region and the largest and the smallest pooled prevalence of antenatal care dropout was reported in a national wide research results, 47.04% (95% CI: 37.86, 56.27) and Tigray region 33.39% (95% CI: 29.45, 37.34) respectively. The low prevalence in the Tigray region might be due to the small sample size of the study used. Additionally, Subgroup analysis was done by year of studies conducted and revealed that studies conducted since 2019 had the highest pooled prevalence of antenatal care dropout 44.35% (95% CI: 34.05, 54.65), compared with studies conducted before 2020. One possible reason could be that the Ethiopian government’s emphasis on addressing the low uptake of maternal health services through various measures to improve antenatal care[25].

Another aim of this systematic review and meta-analysis was to find the most important variables influencing the prevalence of antenatal care service dropout in Ethiopia. Accordingly, distance from the health care facility, marital status, level of education, place residence, pregnancy complication signs and age group) were identified as significant determinants of ANC dropout.

This study found that women who travel more than 1h to reach the health institution was more likely to have antenatal care dropout than their counterparts. This result was supported by studies conducted in Nigeria [26], South Africa [27], southern Ghana [28] and Tanzania [29]. This might be due to the fact that some rural women in developing nations spend more time on their multiple responsibilities, such as care children, gathering water or fuel, cooking, cleaning, and engaging in agricultural work. As a result, they may not be able to travel long distances or use different transportation services to get into the health facilities to access health care services.

This study found that the pregnant women who had no pregnancy complication signs was significantly associated with antenatal care dropout. This study report was supported by a study results done in Hadiya zone Ethiopia [30], South Sudan [31], Malawi [32] and low-income and middle income countries [33]. The possible justification might be due to low attention of health care seeking, and a lack of community access to the health care system.

Furthermore, this meta-analysis revealed that place of residence had significantly associated with antenatal care dropout. Pregnant women from rural areas had ANC dropout than women from urban areas. This finding is consistent with studies conducted in Ethiopia [34], Indonesia [35], Nigeria [29, 36]. The possible justification could be due to the less availability of health care facilities in rural areas. And also this study could suggests that urban women may have better access to health care services and have a better understanding of prenatal care than rural women.

Also, this meta-analysis revealed that women’s educational level had significantly associated with antenatal care dropout. Women who had no educational level were more likely to dropout from ANC compared to those women who have secondary and above educational level. This finding is consistent with studies report conducted in Enemay district Ethiopia [37], Pakistan [38], Nigeria [39], and Egypt [40]. The possible reason could be the direct relationship between women’s education and preference for prenatal care. Those who are more knowledgeable about the importance and appropriateness of ANC services are more likely to take advantage of the recommended number of ANC visits [41].

Finally, women who were in the age group of 30-49 years lower antenatal care dropout than women the age group of 15-29. The possible justification could be due to the fact that women of older age are more able to make their own health care decisions, which in turn increases the likelihood that pregnant women will use ANC services. This study suggest that early age at marriage has a negative impact on ANC utilization.

### Limitations of the Study

Despite the fact that this systematic review and meta-analysis on antenatal care dropout and its associated factors was the first of its kind in Ethiopia, the limitations listed below should be considered when interpreting the results. Due to the nature of the study design, most of the studies considered were cross-sectional, making it difficult to establish a cause-and-effect relationship. In addition, the studies were from three regions, which may restrict the generalizability of the findings. And finally, due to the lack of sufficient literature on this topic, we were forced to compare some of our findings with primary studies conducted aboard.

## Conclusion and recommendations

This meta-analysis revealed the pooled prevalence of antenatal care service among pregnant women 41.37% in Ethiopia. The major determinant factors associated with ANC drop-out were distance from the health facility, place of residence, educational level, pregnancy danger sign and age of women. Hence, to reduce the number of ANC dropouts it is important to counsel and educate women at their first prenatal care. Issues of urban-rural disparity and locations identified as hotspots for incomplete ANC visits require further attention.

## Data Availability

The data underlying the results presented in the study are submited

## Acknowledgements

The authors would like to thank the authors of the studies included in this systematic review and meta-analysis.

## Author Contributions

**Gizaw Sisay:** Conceptualization; formal analysis; methodology; validation; writing the original draft. **Tsion Mulat:** Data curation; methodology; software; writing, review & editing the manuscript

## Ethical approval and consent to participate

Not applicable; since we did not use primary data that needs ethical approval and consent to participate.

## Availability of data and materials

All the datasets used in this study are accessible in the published article and its Additional files.

## Reference

1. Tunçalp, ⍰., et al., WHO recommendations on antenatal care for a positive pregnancy experience-going beyond survival. Bjog, 2017. 124(6): p. 860–862.

2. Bergsjø, P., What is the evidence for the role of antenatal care strategies in the reduction of maternal mortality and morbidity? Safe motherhood strategies: a review of the evidence, 2001.

3. Organization, W.H., Maternal mortality: evidence brief. 2019, World Health Organization.

4. Zolfaghari, E., Z. Boroumandfar, and N. Nekuei, Comparison of reproductive health and its related factors in vulnerable and nonvulnerable women. Journal of Education and Health Promotion, 2022. 11.

5. Muluneh, A.G., et al., High dropout rate from maternity continuum of care after antenatal care booking and its associated factors among reproductive age women in Ethiopia, Evidence from Demographic and Health Survey 2016. PloS one, 2020. 15(6): p. e0234741.

6. Tiruaynet, K. and K.F. Muchie, Determinants of utilization of antenatal care services in Benishangul Gumuz Region, Western Ethiopia: a study based on demographic and health survey. BMC pregnancy and childbirth, 2019. 19(1): p. 1–5.

7. Organization, W.H., WHO recommendations on antenatal care for a positive pregnancy experience. 2016: World Health Organization.

8. Organization, W.H., Trends in maternal mortality 2000 to 2017: estimates by WHO, UNICEF, UNFPA, World Bank Group and the United Nations Population Division. 2019.

9. Jiru, H.D. and E.G. Sendo, Promoting compassionate and respectful maternity care during facility-based delivery in Ethiopia: perspectives of clients and midwives. BMJ open, 2021. 11(10): p. e051220.

10. Mulat, A., et al., Missed antenatal care follow-up and associated factors in Eastern Zone of Tigray, Northern Ethiopia. African Health Sciences, 2020. 20(2): p. 690–696.

11. Chikako, T.U., et al., Multilevel Modelling of the Individual and Regional Level Variability in Predictors of Incomplete Antenatal Care Visit among Women of Reproductive Age in Ethiopia: Classical and Bayesian Approaches. International Journal of Environmental Research and Public Health, 2022. 19(11): p. 6600.

12. Bekele, Y.A., et al., Determinants of antenatal care dropout among mothers who gave birth in the last six months in BAHIR Dar ZURIA WOREDA community; mixed designs. BMC Health Serv Res, 2020. 20(1): p. 846.

13. Mulugeta, B., Antenatal Care Dropout and Associated factors Among Mothers Who Gave Birth in The Last Three Months in North Mecha District, Northwest Ethiopia, 2021. 2021.

14. Tariku, M., et al., More Than One-Third of Pregnant Women in Ethiopia Had Dropped Out From Their ANC Follow-Up: Evidence From the 2019 Ethiopia Mini Demographic and Health Survey. Front Glob Womens Health, 2022. 3: p. 893322.

15. Worku, D., et al., Antenatal care dropout and associated factors among mothers delivering in public health facilities of Dire Dawa Town, Eastern Ethiopia. BMC Pregnancy Childbirth, 2021. 21(1): p. 623.

16. Health, E.F.M.o., Health sector transformation plan I. Adis Ababa, 2015.

17. Page, M.J., et al., The PRISMA 2020 statement: an updated guideline for reporting systematic reviews. Systematic reviews, 2021. 10(1): p. 1–11.

18. Buccheri, R.K. and C. Sharifi, Critical appraisal tools and reporting guidelines for evidence-based practice. Worldviews on Evidence-Based Nursing, 2017. 14(6): p. 463–472.

19. Institute, J.B., JBI Checklist for Analytical Cross Sectional Studies. JBI Global, 2020.

20. Organization, W.H., Trends in maternal mortality: 1990-2015: estimates from WHO, UNICEF, UNFPA, World Bank Group and the United Nations Population Division. 2015: World Health Organization.

21. Akinyemi, J.O., R.F. Afolabi, and O.A. Awolude, Patterns and determinants of dropout from maternity care continuum in Nigeria. BMC pregnancy and childbirth, 2016. 16(1): p. 1–11.

22. Kilowua, L.M. and K.O. Otieno, Health System Factors Affecting Uptake of Antenatal Care by Women of Reproductive Age in Kisumu County, Kenya. International Journal of Public Health and Epidemiology Research, 2019. 5(2): p. 119–124.

23. Francis, W.P., Assessment of Dropout Rate and Contributing Factors among Women Attending Antenatal Care in Samia Bugwe North Busia District: Health Facility Based Survey, June 2013. Research Journal of Medical Science & Public Health, 2015. 2(1): p. 1–10.

24. Singh, D.R. and T. Jha, Exploring factors influencing antenatal care visit dropout at government health facilities of Dhanusha District, Nepal. American Journal of Public Health Research, 2016. 4(5): p. 170–5.

25. Health, E.M.o., Health Sector Transformation Plan II 2020/2021–2024/2025. Ethiop Minist Heal [Internet], 2021. 25(February): p. 1–128.

26. Olayinka, A., A. Joel, and D. Bukola, Factors influencing utilization of antenatal care services among pregnant women in Ife Central Lga, Osun State Nigeria National Hospital Abuja, Nigeria. Advances in Applied Science Research, 2012. 3(3): p. 1309–1315.

27. Worku, E.B. and S.A. Woldesenbet, Factors that influence teenage antenatal care utilization in John Taolo Gaetsewe (JTG) district of northern Cape Province, South Africa: underscoring the need for tackling social determinants of health. International Journal of MCH and AIDS, 2016. 5(2): p. 134.

28. Manyeh, A.K., et al., Factors associated with the timing of antenatal clinic attendance among first-time mothers in rural southern Ghana. BMC pregnancy and childbirth, 2020. 20(1): p. 1–7.

29. Adewuyi, E.O., et al., Prevalence and factors associated with underutilization of antenatal care services in Nigeria: A comparative study of rural and urban residences based on the 2013 Nigeria demographic and health survey. PloS one, 2018. 13(5): p. e0197324.

30. Abosse, Z., M. Woldie, and S. Ololo, Factors influencing antenatal care service utilization in hadiya zone. Ethiopian Journal of Health Sciences, 2010. 20(2).

31. Mugo, N.S., M.J. Dibley, and K.E. Agho, Prevalence and risk factors for non-use of antenatal care visits: analysis of the 2010 South Sudan household survey. BMC pregnancy and childbirth, 2015. 15(1): p. 1–13.

32. Wang, W., et al., The relationship between the health service environment and service utilization: linking population data to health facilities data in Haiti and Malawi. 2015: ICF International Rockville.

33. Kuhnt, J. and S. Vollmer, Antenatal care services and its implications for vital and health outcomes of children: evidence from 193 surveys in 69 low-income and middle-income countries. BMJ open, 2017. 7(11): p. e017122.

34. Tegegne, T.K., et al., Antenatal care use in Ethiopia: a spatial and multilevel analysis. BMC pregnancy and childbirth, 2019. 19(1): p. 1–16.

35. Titaley, C.R., M.J. Dibley, and C.L. Roberts, Factors associated with underutilization of antenatal care services in Indonesia: results of Indonesia Demographic and Health Survey 2002/2003 and 2007. BMC public health, 2010. 10(1): p. 1–10.

36. Babalola, B.I., Determinants of urban-rural differentials of antenatal care utilization in Nigeria. African Population Studies, 2014. 28(3): p. 1263–1273.

37. Shitie, A., et al., Completion and factors associated with maternity continuum of care among mothers who gave birth in the last one year in Enemay district, northwest Ethiopia. Journal of Pregnancy, 2020. 2020.

38. Iqbal, S., et al., Continuum of care in maternal, newborn and child health in Pakistan: analysis of trends and determinants from 2006 to 2012. BMC health services research, 2017. 17(1): p. 1–15.

39. Onasoga, O.A., J.A. Afolayan, and B.D. Oladimeij, Factor’s influencing utilization of antenatal care services among pregnant women in Ife Central LGA, Osun State Nigeria. Advances in Applied Science Research, 2012. 3(3): p. 1309–1315.

40. Hamed, A., E. Mohamed, and M. Sabry, Egyptian status of continuum of care for maternal, newborn, and child health: Sohag Governorate as an example. Int J Med Sci Public Health, 2018. 7(6): p. 1.

41. Ali, S.A., et al., Factors affecting the utilization of antenatal care among pregnant women: a literature review. J Preg Neonatal Med, 2018. 2(2).

